# Transcriptomic analyses reveal regional signatures in lung allograft recipients

**DOI:** 10.1101/2023.04.03.23288063

**Authors:** Patricia Agudelo-Romero, Kak-Ming Ling, Melanie A. Lavender, Jeremy P. Wrobel, Michael Musk, Stephen M Stick, Anthony Kicic

## Abstract

**Background:** Long term outcomes of allograft recipients are compromised by the development of chronic lung allograft dysfunction (CLAD) promoting bronchiolitis obliterans syndrome (BOS). We established baseline transcriptomic profiles of both the large and small airway epithelial cells (referred as LAEC and SAEC, respectively) to identify regional differences irrespective of initiating disease.

**Methods:** We obtained matched primary LAEC and SAEC from lung allograft recipients (n=4, 42.5 ± 4.2 years) and established primary cultures. Bulk RNA sequencing was performed to determine differentially expressed genes.

**Results:** We observed differences in the transcriptional program between LAEC and SAEC Transcription factors (TF) were ranked within the top ten differentially regulated genes. The most abundant TF families included C2H2-ZF, homeobox and bHLH. Upstream regulator analyses identified homeobox genes being significantly in LAEC. Protein-protein interaction network analysis emphasised the role of TFs (*ISL1, MSX1, HOXA1, GATA6, ZNF423*) in airway modulation. Additionally, functional enrichment analysis revealed the activation of chemotaxis, metalloendipeptidase/metallopeptidase activity and pro-inflammatory signatures (IL17 signalling and RAGE), in LAEC, while SAEC were characterised by elevated expression of surfactant metabolism related genes. Moreover, alveolar and club cells-related genes were expressed in SAEC, suggesting a lower airway-specific signature.

**Conclusion:** Our analysis shows robust transcriptional differences between LAEC and SAEC. We suggest a potential role for homeobox TF family as well as the activation of the immune system in the biology of LAEC. Conversely, we observed an alveoli-like transcriptional signature in SAEC, including gas-exchange signals and surfactant metabolism; pathways involved in lung homeostasis.

## Introduction

Chronic allograft dysfunction (CLAD) due to bronchiolitis obliterans syndrome (BOS) is a major complication following pulmonary transplantation whose aetiology is not fully understood^1^. Recent findings indicate that the pathology of CLAD is not only limited to the small airways, but also manifests in the proximal airways^2^. The underlying mechanism of BOS is suggested to involve injury and inflammation of epithelial and subepithelial cells, which in turn stimulates wound repair and epithelial to mesenchymal transition signaling^3^. Thus, understanding the pathological mechanisms contributing to the onset of BOS, could facilitate tailored interventions aimed at preventing this pathology.

For the purpose of tracking the different processes that might lead to the onset of BOS, we conducted a baseline study, to compare matched samples of proximal and distal lung airways, without any sign of disease. We did RNA-seq to explore regional transcriptional differences in epithelial cultures established from large/proximal and small/distal airway epithelial cells (referred as LAEC and SAEC) from Lung Transplant recipients. Here, we observed that distal airways culture expressed genes related to surfactant metabolism. Whereas proximal airways cultures displaying two proinflammatory pathways and fibrosis pathways that might be related to early dysregulation of BOS; providing evidence that alterations in gene expression begin in the proximal airways.

## Materials and Methods

### Patient and sampling procedures

Matched samples from proximal and distal lung biopsies were obtained from lung allograft recipients during the routine surveillance bronchoscopy (Figure S1A). Patients did not present symptomatology indicative of bronchiolitis obliterans syndrome (BOS), transplant rejection or infections at the time of sample collection. Specimens were obtained from four patients (3 females, age range 38-47) that underwent double lung transplantation as previously described^4^. The reasons for lung transplantation were cystic fibrosis (3 cases), and congenital heart disease (1 case). Both, LAEC and SAEC were grown as a monolayer to confluency and cells then harvested for transcriptomics analysis (Figure S1B). The study was approved by the Royal Perth Hospital, Ethics Committee (Registration: EC2006/021) and written consent was obtained from each participant after being fully informed about the premise and purpose of the study. A summary that outlines the analyses performed here is presented in Figure S2.

### Total RNA extraction, library preparation and sequencing

Total RNA was extracted using the PureLink® RNA kit (Life Technologies) following their instructions and treated with RNase Inhibitor (Applied Biosystems™) and DNase I (Invitrogen™). RNA concentration and integrity were then determined using a NanoDrop system and an Agilent Bioanalyser, respectively. Libraries were generated using TruSeq Stranded mRNA (Illumina) kit and Poly(A) enrichment. Sequencing was performed using 100 bp single-end configuration (SE; 100 bp; 20 M).

### Bioinformatic and statistical analyses

FASTQ files were quality controlled using FastQC (https://www.bioinformatics.babraham.ac.uk/projects/fastqc/). Reads were trimmed and adapters removed with Trimmomatic^5^ (Figure S3A and S3B). High-quality reads were then mapped to human reference genome (hg19/GRCh37, Ensembl) using Spliced Transcripts Alignment to a Reference (STAR)^6^. Gene-level quantification of raw counts was performed using High-Throughput Sequencing^7^ (HTSeq; Supplementary Materials S1) and post-alignment stats were generated with MultiQC^8^ (Figure S3C and S3D). Gene expression analysis was done using EdgeR^9^ and limma^10^. Latent variance was treated with removeBatchEffect function from limma^10^. Expression values were used for cluster analysis using pvclust R package^11^ (Figure S4; Supplementary Materials S2). *P*-values were corrected for multiple-testing using the Benjamini-Hochberg’s method, False Discovery Rate (FDR)^12^. Ranking metric for top regulated genes was calculated as follow: [Rank = -sign(log_2_FC) * log_10_(FDR)]. Differentially expressed genes (DEGs) were then filtered considering an FDR ≤0.05 and a Fold-Chance (FC) ≥|1.5| and annotation was added using the org.Hs.eg.db R package^13^ (Supplementary Materials S3). Volcano plot was generated using EnhancedVolcano R package^14^.

Mining of transcription Factors (TF) in our data was done following the pipeline described by Lambert *et al*.,^15^ (Supplementary Materials S4) Analysis of upstream transcriptional regulators was performed with ChEA3^16^ (Supplementary Materials S5). Protein-protein interaction networks were generated using NetworkAnalyst^17^ and subsequently analysed using NetworkAnalyzer App with NetworkAnalyzer App (Supplementary Materials S6). Enrichment analyses of gene ontology (GO), KEEG and Reactome databases were done using ClusterProfiler^18^ (Supplementary Materials S7). The RNA-sequencing data have been deposited in the National Center for Biotechnology Information’s Bioproject PRJNA885405 and Gene Expression Omnibus (GEO) under accession number GSE214434 (https://www.ncbi.nlm.nih.gov/geo/query/acc.cgi?acc=GSE214434).

## Results

To determine any regional transcriptional differences between the lung airways, LAEC were compared to SAEC. Hereafter, up-regulated genes will be referred as active in LAEC and down-regulated genes as active in SAEC.

### Gene expression differences between lung regional comparison

After filtering, a total of 355 differentially expressed genes (DEGs), 188 up-regulated and 167 down-regulated genes were identified (Figure 1A). A volcano plot shows the bidirectional distribution patterns of up- and down-regulated genes along with the top genes (Figure 1B). Among the top 10 ranked genes, two transcription factors (TFs) (*ZFHX4* and *MSX1*) belonging to homeobox family were found to be up-regulated, and *ZNF730* belonging to C2H2 Zinc Finger (ZF) which was down-regulated.

**Figure 1.**
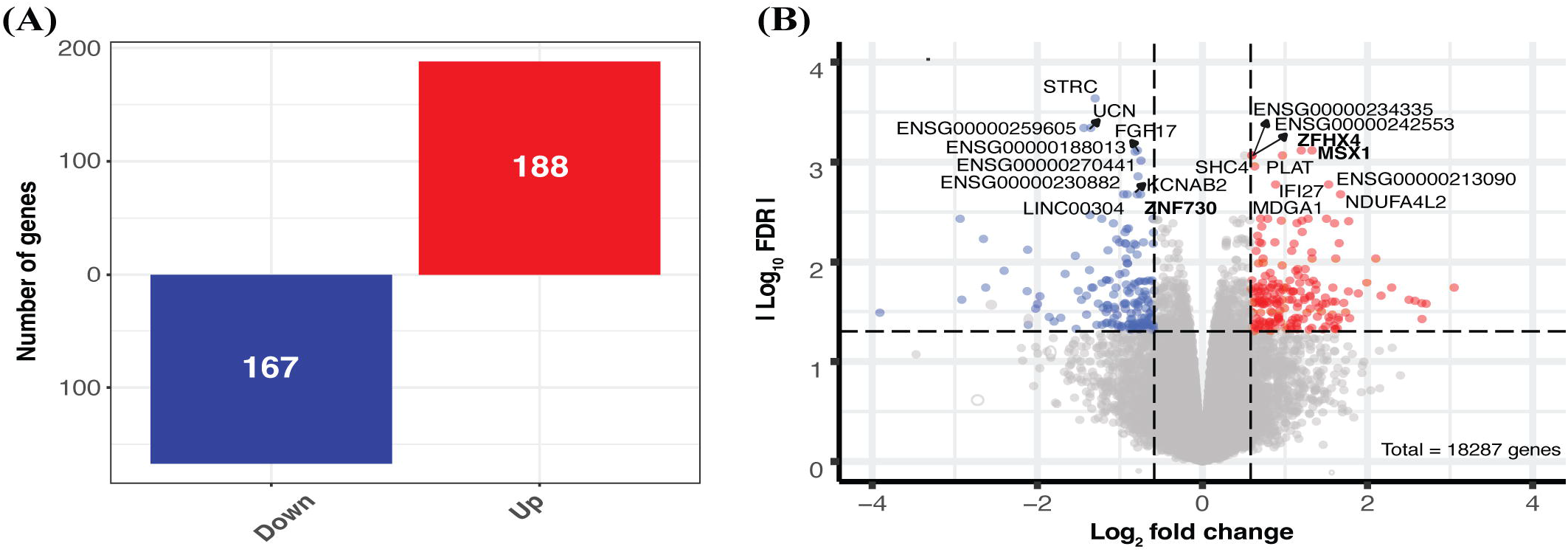
Transcriptomic signatures between proximal and distal lung regions. (A) Bar plot summarises the number of DEGs (FC ≥ |1.5| and FDR < 0,05) of the regional comparison on the y-axis and direction of the change on the x-axis, divided in up- and down-regulated genes, up (red) and down (blue), respectively. (B) Volcano plot displays bidirectional log2 FC on the x-axis and absolute Log10 FDR on the y-axis. Top 10 ranked genes for up- and down-regulated genes are labelled in each case and bold letter highlight Transcription Factor (TF) genes.

### Transcript Factors modulation underlies gene expression differences between lung airways

Knowing the essential role TF have in gene modulation, we explored their potential contribution towards the differences seen between the LAEC and SAEC gene transcriptional profiles. For this purpose we made used of the human TFs catalogue^15^. We detected a total of 26 TF belonging to eight TF families and the three most abundant including C2H2 ZF (34.6 %), homeobox (26.9 %) and bHLH (11.5 %) families (Figure 2A; Supplementary Materials S4). Using Venn diagram analysis, two overlapping families were identified from up- and down-regulated genes comparison (C2H2 ZF and homeobox) (Figure 2B). More interesting, four TFs families were uniquely identified in LAEC (BED ZF, DM, Fox and T-box), and two specifics in SAEC (bHLH and GATA) (Figure 2B). Bar plots summarise the 14 up- and 12 down-regulated TFs (Figure 2C).

**Figure 2.**
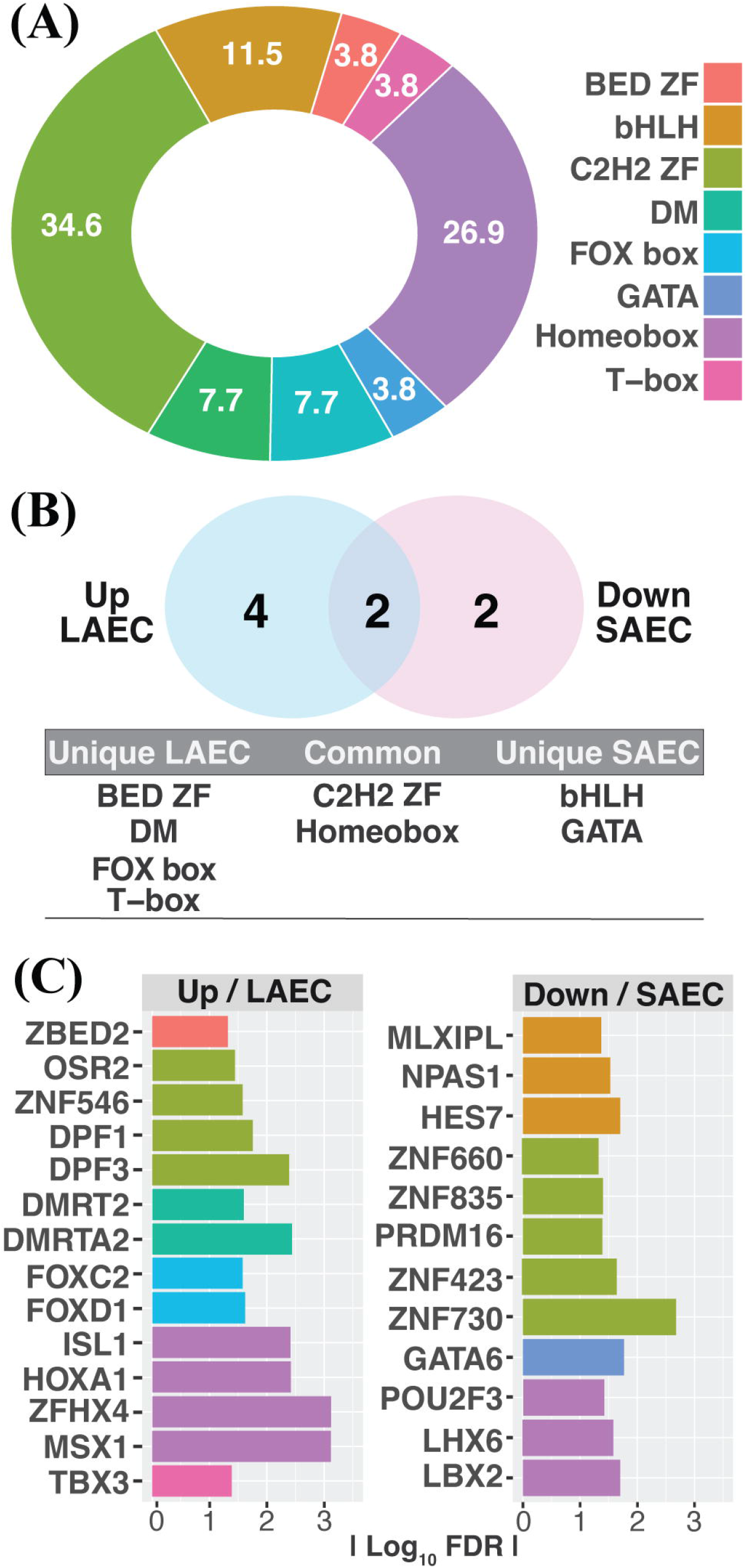
Identification of Transcription Factors (TF) regulating proximal and distal lung regions. (A) Pie plot represents the percentage of TF families from the TF in DEGs. (B) Venn diagram highlights the common and uniquely TF families between LEAC and SAEC. Blue and pink represent proximal/LAEC and distal/SAEC samples. (C) Bar plots depict the TF signature found between up- and down-regulated as LAEC and SAEC, respectively. Colour palette refers to the TF families.

### Homeobox genes represent central modulators of gene expression in the proximal airways

To gain insight into the gene regulatory network of the airways and identify upstream modulators linked to the observed DEGs, we performed a TF enrichment analysis (TFA). We retrieved 161 upstream regulators significantly associated with the up-regulated genes, of which eight TF belonging to homeobox family were observed in the top ten upstream regulators (Figure 3A). In contrast, no upstream regulators for the down-regulated genes were identified. To better understand the interaction between TFs and modulated genes, we then generated networks based on the relevant associations (Figure 3B; Supplementary Materials S5). Interaction between six upstream regulators explained the activation of 18 genes. *HOXD10* and *HOXD9* represented central modules explaining differences in gene expression of five genes, three of which are TF (*MSX1, TBX3* and *FOXD1*) (Figure 3B). Lastly, topological analyses were used to identify hub genes from the DEGs. This network analysis employed 823 nodes with at least one connected component, where 97 annotated genes belong to our DEG core. Hub genes were extracted by filtering nodes above 14 degrees, which resulted in a set of 24 genes (17 up-regulated and 7 down-regulated genes) (Figure 3C) and the top five hubs are *SPP1, HOXA1, S100A8, KRT14* and *ID2*. Besides, several TFs were identified as hub genes, three up-regulated (*ISL1*; *MSX1*; *HOXA1*) and two down-regulated (*GATA6*; *ZNF423*). Again, the three hub TFs from up-regulated genes belong to homeobox family.

**Figure 3.**
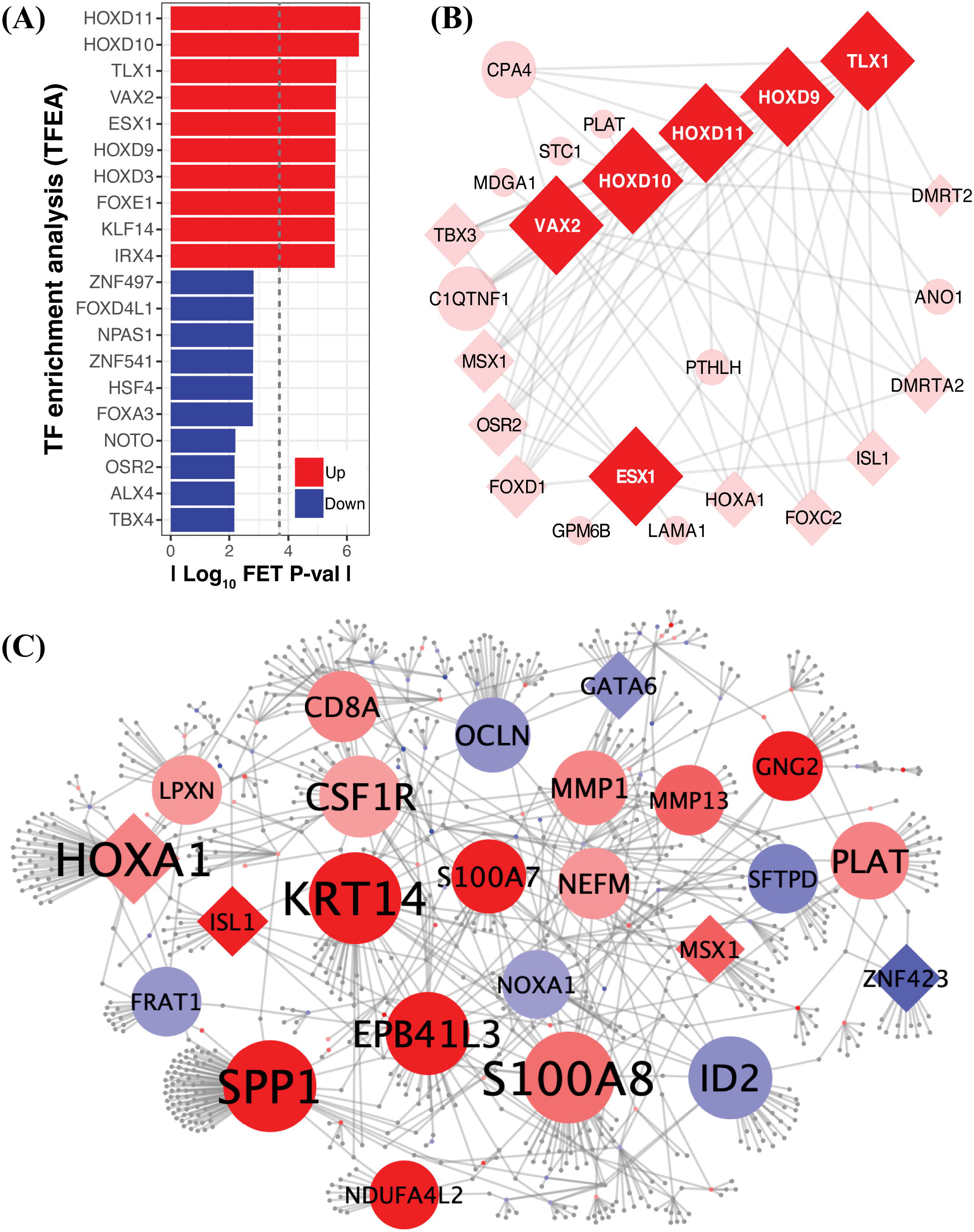
Upstream regulator enrichment and transcriptional network analyses of significantly differentially expressed genes. (A) Bar plot visualisation of TF enrichment analysis (TFEA) shows the top upstream regulators enriched from the DEGs core using a Fisher’s exact test (FET) from ChIP-X Enrichment Analysis 3 (ChEA3)16. Up- and down-regulated genes are represented in red and blue, respectively. Dash line represents the FDR 5% limit. (B) Network analysis of upstream regulators and associated regulated genes from up-regulated genes. Network was clustered using an Edge-weighted Spring-Embedded Layout method46. Brighter colours illustrate upstream regulators. Shapes depict genes and TF as circles and diamonds, respectively. (C) Protein-protein interaction network of the differentially expressed genes (DEG) was calculated from IMEx Interactome database of InnateDB (https://www.innatedb.com) using NetworkAnalyst17, it was analysed and visualised in Cytoscape46. Colour intensity displays the fold change value (red; up-regulated genes and blue; down-regulated genes), edges are show in light grey; beside node size represents the degree centrality as interconnection.

### Functional enrichment shows a pro-inflammatory signature in proximal airway and pulmonary surfactant metabolism in the distal airway

We next performed functional enrichment to determine the biological pathways associated with the up- and down-regulated genes. Specifically, we utilised pathway over-representation analysis using GO, Reactome and KEGG databases followed by a category-gene network (Figure 4A-B, Supplementary Materials S7). Several categories were identified enriched from the up- and down-regulated gene list. The top two more statistically significant pathways were taxis/chemotaxis and activity of metalloendipeptidase/metallopeptidase. Besides two pro-inflammatory pathways (RAGE receptor biding and IL17 signalling pathways) were also enriched. Conversely, two pathways representing the down-regulated genes were observed: benzaldehyde dehydrogenase activity and surfactant metabolism.

**Figure 4.**
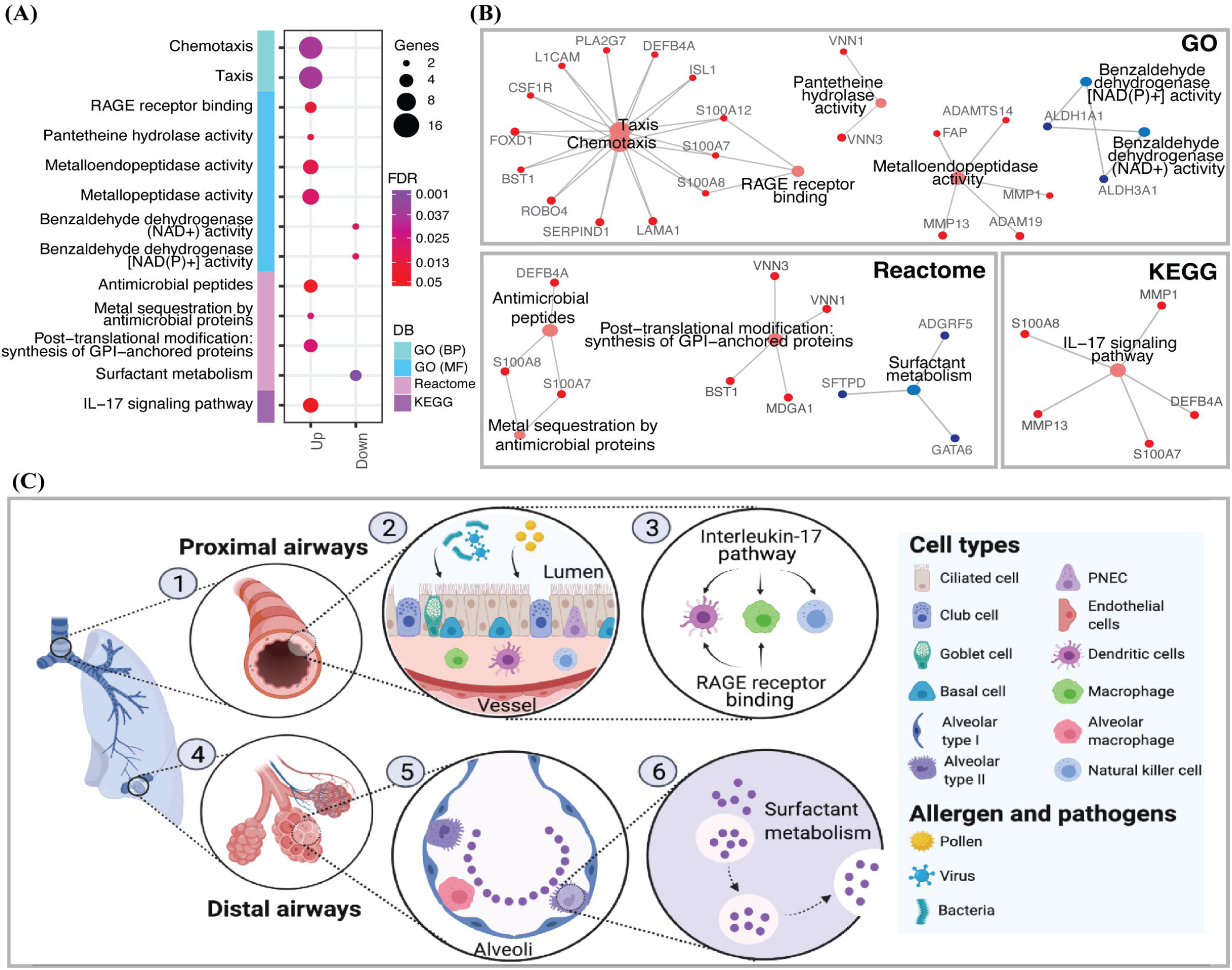
Functional enrichment analyses and transcriptional hallmarks of the proximal and distal airways from lung transplant (LT*x*) recipients. (A) Scatter plot represents the significant categories enriched in up- and down-regulated genes. Bar side row represents databases used (DB), adjusted P-value (False Discovery Rate, FDR) colours the significance and dot size the numbers of genes involved in each category. (B) Category-gene network associations of enriched terms. Up-regulated genes in red and down-regulated genes in blue. (C) Transcriptional hallmarks overview of the airways. Samples from the proximal airways were collected close to the trachea (1). This region is constantly exposed to abiotic (allergens, cigarettes) and biotic (virus, bacteria, fungi) factors (2). Two proinflammatory pathways (IL-17 signalling and RAGE receptor binding) associated with defence mechanisms were found activated in this region (3). Distal airways specimens were collected near the alveoli (4), where the gas-exchange occurs (5). In agreement, these samples demonstrated abundance of transcripts related to surfactant metabolism, which is crucial to lower the alveolar surface tension during expiration (6). Figure created with BioRender.com.

## Discussion

The study of the transcriptional lung regional differences in lung allograft recipients provides an excellent tool to understand baseline molecular mechanisms/signatures that may drive chronic allograft rejection. With this objective in mind, we analysed transcriptional differences between LAEC and SAEC from lung allograft recipients. Our results support the hypothesis that airway transcriptional regional differences exist, where TF genes, specifically homeobox TF may have a potential role in establishing these differences. Protein-protein interaction network analysis emphasised the role of *ISL1, MSX1, HOXA1* in LAEC, and *GATA6, ZNF423* in SAEC. Furthermore, functional enrichment analysis identified activation of chemotaxis, metalloendipeptidase/metallopeptidase and two pro-inflammatory categories in DEGs from LAEC and surfactant metabolism in SAEC DEGs.

Several TF (∼7%) were noticed in the top ten genes of the DEG (Figure 1). Detailed analysis about TF families’ composition in the airways found homeobox and C2H2 ZF in common (Figure 2). Homeobox is related with patterning in lung branching^19^, and although C2H2 ZF plays important roles in development and disease, still remain poorly characterized^20^. Four unique TF families were up-regulated, including BED-ZF, DM, FOX, and T-box, which are involved in development, among other functions. Several TF identified in these families serve primary roles in the lung. For instance, *ZBED2* has been predicted to promote the keratinocyte basal state, inducing differentiation^21^. In addition, *DMRT2* is involved in establishing left–right asymmetry and somitogenesis; whereas *DMRTA2*, also known as *DMRT5*, has been reported in anterior neural tissue development ^22^. Moreover, *FOXD1* has been linked as a marker for lung pericytes and *FOXC2* in vascularisation^23^. Finally, *TBX3* has been found to be involved in lung branching morphogenesis with expression in the lung mesenchyme ^24^. In contrast, two specific TF families, bHLH and GATA, were identified in the down-regulated genes. The bHLH family is important in regulating embryonic development, and *NPAS1* has been reported to regulate branching morphogenesis in the embryogenic lung^25^. Furthermore, the GATA family plays an important role in both epithelial and smooth muscle cell linage diversity in the lung. Specifically, *GATA6* has been found to induce differentiation of primitive foregut endoderm into respiratory epithelial cell linages, in addition to regulating surfactant protein genes^19^.

Next, upstream regulator enrichment analysis of the up-regulated genes predicted that *HOXD9* and *HOXD10* homeobox genes activate *MSX1, TBX3* and *FOXD1* (Figure 3B). Interestingly, *MSX1* and *TBX3* are expressed in mesenchyme cells in the single cell (sc) lung map website^26^ (LungGENS, 10X sc 24-years-old), and have been shown to be regulated by Hedgehog and Wnt signalling pathways^24,27^. Network analyses identified 24 hub genes (Figure 3C); from the up-regulated hubs we found a conserved signature (*CSF1R*; *MMP1*; *CD8A*; *PLAT*; *S100A8*; *SPP1*) similar to that of the tracheal / proximal airway^28^, with genes involved in a variety of processes including cell migration, inflammation response and chemokine production. Moreover, the three TF detected in the up-regulated gene hubs are related to morphogenesis; two specific to mesenchyme development and the other to proximal airways development^27,29^.

Interestingly, several pathways were statically significative in a functional enrichment analysis (Figure 4). From the up-regulated gene list, taxis/chemotaxis processes had the most statistical significative values. Chemotaxis, defined as the directed migration of cells towards a specified entity, are important for cellular pattering and development^30^. In drosophila, chemotaxis has been detected in the tracheal epithelium^31^. Likewise, the metalloendipeptidase activity pathway was also observed to be activated through two matrix metalloproteinase (MMP) genes (*MMP1, MMP13*). In skin repair, *MMP1* alters the migratory substratum driving the forward movement of the repairing cells by allowing them to attach, dislodge, then reattach to the wounded matrix^32^. Interestingly, *MM13* has been shown to play a role in the pathogenesis of liver and lung fibrosis ^33–35^. Additionally, two proinflammatory (IL17 signalling and RAGE receptor binding) plus an antimicrobial pathway were enriched in the up-regulated genes. Amongst these genes, *S100A7* and *S100A8* are known to expressed in the trachea^36^ and have been reported to have a role in innate immune responses to pathogens^37^. Both have gained research interest because they exhibit selectivity towards pathogenic bacteria, while having no effect on beneficial commensal bacteria^38,39^.

Interestingly, the up-regulated enriched categories are also related to epithelial repair and innate immunity and may suggest that their activation after lung transplantation may help prevent the entry of undesirable microorganisms via the production of chemokines and antimicrobial peptides^32^. The inflammation category activation also supports this hypothesis, since it can act as a defence mechanism to abiotic^40^ and biotic^41^ insults. However, a fibrosis-related gene was also observed within the LAEC signature, which in the long term could function to compromise lung allograft outcomes^33–35^.

Two hub upstream regulators from the up-regulated genes, *HOXD9* and *HOXD10*, are known to activate TF involved with the chemotaxis process (*FOXD1*; *ISL1*) (Fig 3B and 4B) and genes related to mesenchymal cells (*ISL1; MSX1; TBX3*). In adults, directional movement of chemotaxis modulates different responses depending on whether it is infectious or injurious. In the first, immune cells would be activated and, in the latter, wound healing and tissue regeneration responses are activated. The latter is provided by the regional stem cells that move into damaged areas, produce connective tissue and maintain tissue homeostasis^30^. It is unclear whether this proximal airway signature from LT*x* patients could be due as part of healing/regeneration response after the lung transplantation, or as part of the immune response linked with the proinflammatory processes previously reported^30,42^. Conversely, only two pathways were enriched from the down-regulated gene list. The first, surfactant metabolism, is essential in small airways because it regulated alveolar surface tension and plays a role in protection against oxidants and infection^43^. The second, NAD- and NADP-dependent benzaldehyde dehydrogenase (ALDH) are involved in detoxification and prior work has reported *ALDHA1* and *ALDH3A1* to be involved in the host defence response to toxins in smokers^44^.

We acknowledge a number of limitations to this research. Firstly, due to the precious nature of the samples involved, this pilot study has a limited number of biological replicates from LT*x* recipients. Despite this, there was sufficient sensitivity to obtain DEG using the rigorous analysis pipeline outlined. Also, the limited expansion potential of primary AEC is a limitation^45^ and its effects remain unknown at the transcriptomic level. Nevertheless, we believe that the utilisation of ‘unaltered primary airway cells’ are a significant strength of this study. Likewise, monolayer cultures may oversimplify the multicellular interactions, but a robust and repeatable model with low methodological variation is important. All together, we are confident that limitations are minor and that our results provide new insight into the transcriptional regional differences. For prospective studies, these limitations could be solved with the use cell culture of specialized cells.

In conclusion, the data presented here suggests distinctive signatures from LAECs and SAECs at a transcriptomic level, establishing new insights into regional gene expression in the airways of lung transplant recipients (Figure 4C). Proximal airway gene expression changes included a proinflammatory signature which may indicate a defence mechanism since it is the first barrier of defence, as well as a fibrotic signature which may initiate downstream complications such as BOS establishment. In contrast, the small airways reflect a characteristic alveoli’s hallmark, including gas-exchange signals and secretion of pulmonary surfactant proteins. These results will seed future large scale works to determine the predictive value of these genes as potential biomarkers aimed at maintaining baseline lung health of allograft recipients.

## Data Availability

Raw datasets have been uploaded to GEO, with accession number GSE214434.

https://www.ncbi.nlm.nih.gov/geo/query/acc.cgi?acc=GSE214434

## Data Availability Statement

Raw datasets have been uploaded to GEO, with accession number GSE214434.

## Ethics Statement

The study was approved by the Royal Perth Hospital, Ethics Committee (Registration: EC2006/021) and written consent was obtained from each participant after being fully informed about the premise and purpose of the study.

## Author Contributions

Conceptualization: SMS, AK and PAR. Data curation: PAR. Formal analysis: PAR. Funding acquisition: SMS and AK. Investigation: PAR, KML, MAL, JPW, MM, SMS and AK. Methodology: PAR, KML, MAL, JPW, MM, SMS and AK. Resources: PAR. Visualization: PAR. Manuscript writing: PAR and AK. Manuscript review: PAR, KML, MAL, JPW, MM, SMS and AK.

## Acknowledgments

This work was supported by grants from the McCusker Foundation and the Heart and Lung Transplant Foundation of Western Australia. A.K. is a Rothwell Family Fellow. S.M.S. is an NHMRC Practitioner Fellow. Analysis was performed in the Pawsey Supercomputing Research Centre with funding from the Australian Government and the Government of Western Australia.

## Supplementary Material

1. **Supplementary Material S1**. Raw counts generated with HTSeq^4^, and obtained from an alignment using STAR^3^ to the Ensembl human reference genome hg19/GRCh37.
2. **Supplementary Material S2**. Expression values used for cluster analysis and gene expression profiles.
3. **Supplementary Material S3**. List of 355 differentially expressed genes with a cut-off of FC > |1.5| and FDR of 5%.
4. **Supplementary Material S4**. Data mining of transcript factor families based on Lambert et al.,^12^.
5. **Supplementary Material S5**. Transcription factor enrichment analysis of the upstream regulators using ChEA3^13^.
6. **Supplementary Material S6**. Network analysis of differentially expressed genes generated by Cystoscope^15^. Statistics related to (A) nodes and (B) edges, as well as (C) the subnetwork built in Figure 3C.
7. **Supplementary Material S7**. Enrichment analysis of differentially expressed genes using ClusterProfiler^17^.

