## Supplementary Figures for "Transcriptomic analyses reveal regional signatures in lung allograft recipients"

**Figure S1.** *Experimental design flowchart.*

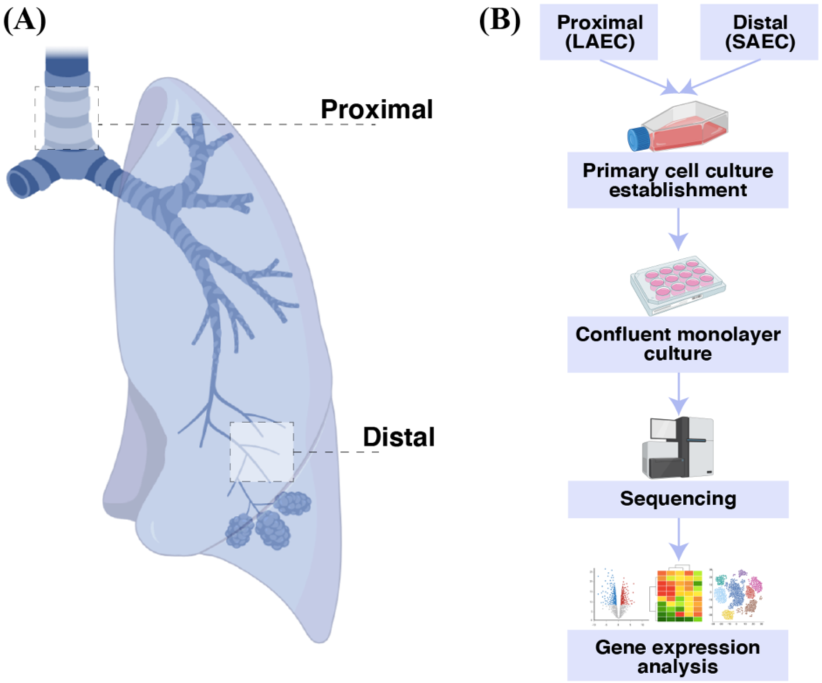


Matched proximal and distal samples were obtained from lung allograft recipients during their routine surveillance bronchoscopy (**Figure S1A**). Primary airway cells of proximal/large and distal/small airway epithelial cells (referred as LAEC and SAEC, respectively) were established and then grown as confluent monolayer culture. Then, samples were collected for RNA sequencing and gene expression analysis (**Figure S1B**). Figure created with BioRender.com.

**Figure S2**. *Map of the gene expression analyses performed through this present study.*

**Methodology**

Sampling and experimental design Fig. S1.

Summary of the analyses performed in the presented study Fig. S2.

Quality control steps of the bioinformatic pipeline Fig. S3.

Table of the raw counts obtained from the alignment Appendix S1.

Clustering analysis of the replicates performed with the expression values Fig. S4 and Appendix S2.

**Hypothesis**

There are transcriptional differences between proximal and distal airways.

**Results**

**Which key genes are differentially expressed between proximal and distal airways?**

- List of differentially expressed genes (DEGs) Appendix S3.
- Number of up- and down-regulated genes Fig. 1A.
- Identification of genes with higher impact in the airway modulation (Top 10 ranked genes) Fig. 1B.

**Which transcription factors (TF) are involved in the regional airways differences?**

- Data mining of the TF families in the DEGs Fig. 2A and Appendix S4.
- Overlapping of the TF families between up- and down-regulated genes Fig. 2B.
- Summary of transcription factor genes detected in the up- and down-regulated Fig. 2C.

**Which upstream regulators are activated in the up- and down-regulated gene lists?**

- Upstream regulator enrichment Fig. 3A and Appendix S5.
- Network of upstream regulator and activated genes from the up-regulated genes Fig. 3B.
- Network of upstream regulator and activated genes from the down-regulated genes Fig. 3C.

**How the differentially expressed genes are organized?**

- Transcriptional network analyses of most interconnected genes Fig. 3D and Appendix S6.
- Biological pathways associated to the differentially expressed genes Fig. 4A and Appendix S7.
- Identification of genes that define cellular identity in the airways Fig. S5 and Appendix S8.

**Conclusions**

Overview of the main regional transcriptional differences identified in lung allograft recipients Fig. 4C.

**Figure S3**. *Quality Control from pre- and post- alignment.*

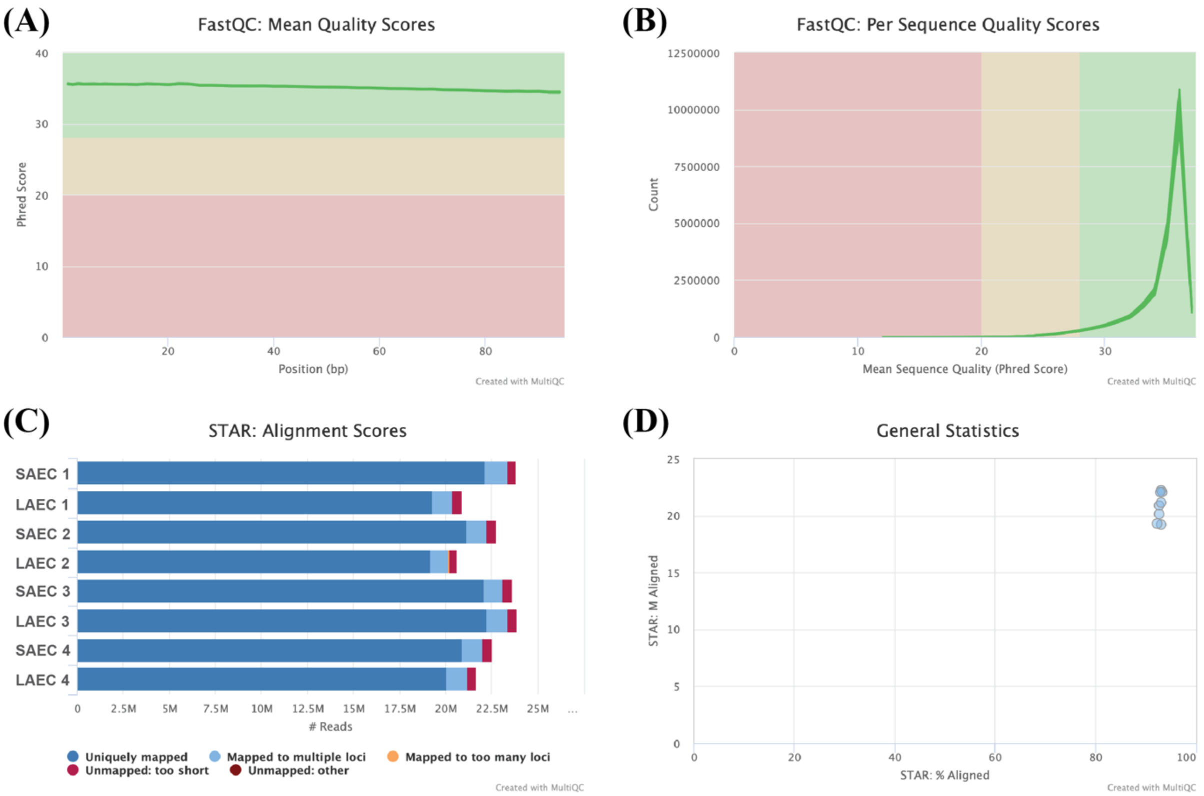


Quality scores plots show Phred scores with the highest values in green (Phred scores > 35), through reads (100bp; **Figure S3A**) and total number of reads (**Figure S3B**). Bar plot of the alignment shows sequences were uniquely mapped (blue) to human genome (**Figure S3C**). Scatter plot depicts the general statistics of aligned reads percentage against reads in Million (**Figure S3D**), where bam files contain ~20 M and ~93 % of the reads were mapped.

**Figure S4.** *Cluster analysis built through a hierarchical dendrogram approach.*

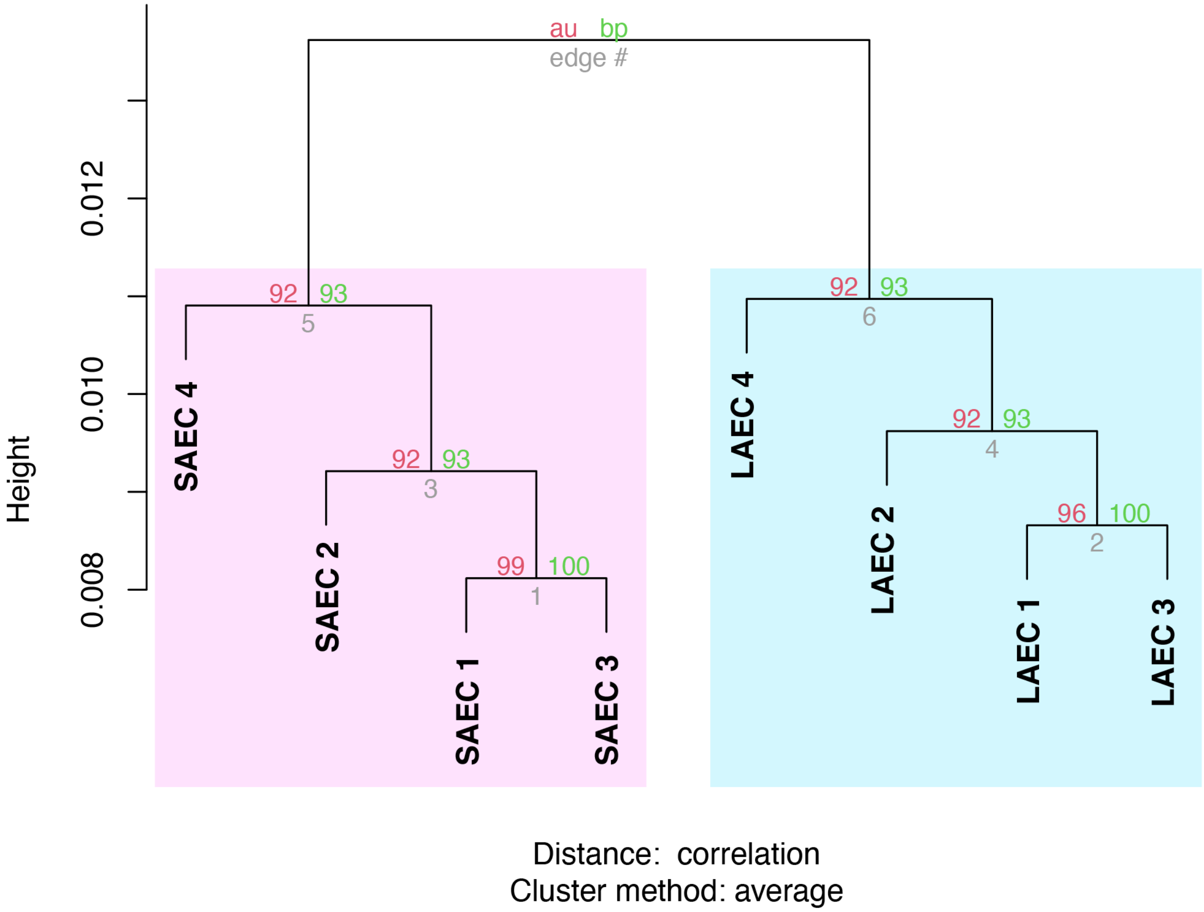


Clustering of replicates by airway region was assessed using a hierarchical clustering via multiscale bootstrap resampling (300 bootstrap), employing parameters "correlation" for distance measure method and "average" for the agglomerative method. Samples are highlighted in pink for distal/SAEC and blue for proximal/LAEC samples (Appendix S2).
